# Mitigating Covid-19 aerosol infection risk in school buildings: the role of natural ventilation, classroom volume, occupancy

**DOI:** 10.1101/2021.03.23.21253503

**Authors:** Alessandro Zivelonghi, Massimo Lai

## Abstract

Issues linked to aerosol physics within school buildings and related infection risk still lack a proper recognition in school safety regulations. In this theoretical work we try to shed more light on the critical role of air ventilation, classroom volume, occupancy, and face mask types (surgical vs ffp2) in controlling airborne transmission risk in educational settings. Limited spaces available in many schools require to precisely assess the occupancy/volume ratio in each classroom and to investigate the specific risk levels from aerosolization of viral loads from infective sources. Moreover, most schools are not provided with mechanical HVAC systems. Fundamental questions are therefore: how the specific classroom volume affects the long-range contagion risk in a given classroom? is linear social distancing the right way to assess a volumetric risk problem? How effective are other countermeasures like reduced speaking volume or equipping teachers with microphones? We present here the results of a numerical analysis based on the Gammaitoni-Nucci infection risk model and the consolidated Wells-Riley like approach, with SARS-CoV2 input data and geometric data from a typical high-school classroom in Italy. We investigated separately the case of infective asymptomatic student and infective asymptomatic teacher as source of viral quanta, taking into account thermal gradient effects on the air ventilation rates. First recommendations based on the volumetric nature of aerosol risk are suggested to extend the linear social distancing approach applied so far. Finally we discuss the concept of “cumulative infection risk” over multiple lesson+break cycles in Wells-Riley-like infection models. We believe that any attempt to proper model infection risk in closed environments with cycled changes of the source and the susceptible individuals, should carefully consider this point, particularly when modelling air ventilation breaks in classrooms.

## 1. Introduction

School classrooms are enclosed settings where students and teachers spend prolonged periods of time and therefore risky environments for airborne transmission of SARS-CoV2. Airborne infections originate from viral aerosol formation and the cumulative nature of air saturation. As stated by Morawska et al. in [1] and recently recognized by WHO, “inhaling small airborne droplets is a probable third route of infection” in addition to transmission via larger respiratory droplets and direct contact with infected people or contaminated surfaces. Evidence of airborne transmission causing outbreaks in different enclosed environments was reported from the early stage of Covid-19 pandemics [2]. Outbreaks in schools have also been reported in different countries from the beginning of pandemics [3-5, 29], albeit the definition of outbreaks may vary. In a study on Israeli schools based on extended slub-testing, however, the occurrence of airborne transmission as probable main cause of infection in crowded classes has been well documented [6].

In a school classroom, groups of students, usually between twenty and thirty individuals, share the same premises for hours with potentially insufficient ventilation. This increases the likelihood of coming into contact with virus-loaded aerosol droplets generated by one infective source (student or teacher). This issue is of concern also when social distancing is correctly implemented because of the volumetric and cumulative nature of aerosol clouds. The hypothetical scenario of an infective asymptomatic source entering a school classroom should be carefully investigated for the potentially large consequences it carries and to define a comprehensive risk mitigation strategy. This approach is valid even if preventive countermeasures are applied to reduce the entrance probability of infective sources. In fact, this probability cannot be curbed to zero, particularly in densely populated areas, where higher density has been shown to correlate to an increase in epidemic curves [11]. As a further evidence, many school outbreaks were recently reported in regions where preventive quarantine was in force after school reopening, like e.g. in Veneto Region, Italy at the end of April 2021 [29].

Indirect oral transmission of SARS-CoV2 is believed to be effectively reduced through the frequent opening of windows or by mechanical HVAC systems [8,27]. Proper ventilation has already been proven to significantly lower oral transmission of other diseases like tuberculosis and influenza in confined environments (e.g., [7-9]). Very recent (although still unpublished) measurements of SARS-CoV2 air concentrations in ventilated and non-ventilated hospital rooms performed by the Italian Regional Environmental Agency “ARPA Piemonte” further confirm this believe for the covid-19 case [10].

Besides natural ventilation, mechanical ventilation systems, when adequately configured, could be equally or even more effective in mitigating the aerosol diffusion [8].

Unfortunately, unlike hospitals, the vast majority of schools worldwide are not equipped with such systems and will not be, at least for the foreseeable future (including the 2020/2021 school year). The present study presents results through simulation of windows opening combined with other mitigations, but the model within certain approximations could be easily adapted to account for specific ventilation levels of HVAC systems.

A comprehensive mitigation strategy is presented here which include the effect of splitting class groups as well as the impact of voice reduction on the aerosol infection risk. Furthermore, we improved the GN-model introducing the indoor/outdoor thermal gradient affecting the airflow during windows opening and the effective volume of dilution to account for a more realistic aerosol physics. Preliminary results by one of the authors [12] were limited to the case of an infective student source without temperature influence on the air exchange rates. In the present work we also highlight the concept of cumulative risk over multiple lesson+break cycles and investigate the separate case of infective teacher and its important implications on the airborne risk mitigation strategy.

## 2. Methods

### 2.1 Gammaitoni-Nucci model with thermal gradient airflow

The infection risk model used in the present analysis implements a Wells-Riley like approach [13] extended in the Gammaitoni-Nucci (GN) model [14] which considers time evolution of the viral charge. These models have been proven effective for measles, tubercolosi and influenza and are adequate for confined and ventilated environments. They are based on the assumption that newly-produced viral particles are instantly diluted over the whole environment volume (perfect-mixing) and that the emission rate parameter ER_q_ (emission rate, i.e. number of viral particles generated per hour by each infective subject) is known, at least as an averaged value over the exposure time 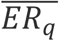 recent paper, authored by several experts in the field [1], elucidated how a possible mechanism for transmission of SARS-Covid2 in confined spaces would be the formation of “light” aerosol droplets (i.e. < 5 µm in diameter, unlike “heavy” droplets, over 5 µm) that diffuse in the environment after being produced by an infected person. In the GN model, if the number of infective sources remains constant, the probability of infection for each subject at a given time *t* will only depend on the total concentration of viral particles supposed isotropically distributed in the volume *V*. This probability follows a well-known exponential law for increasing exposure time t, which strictly depends on the parameter 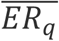 and the ventilation ratio *p/AER*, where *p* is the average inhalation flow (related to pulmonary capacity) and *AER* is the inflow of clean air provided by natural or mechanical ventilation. To account for the complex phenomenon of viral inactivation and gravitational deposition on surfaces [15], the air exchange rate (AER) in (2-4) is more properly substituted from an *infective virus removal rate (IVRR* = *AER* + *λ* + *k)* which adds to the AER a viral inactivation factor (λ) and a particle deposition factor (k). For the purpose of this demonstration the small contribution of particle deposition on surfaces has been neglected also based on the fact that in winter heating systems in a classroom would tend to move air upward. In addition, k values for standard non-heated environments are found in literature below 0.25 vol/h [16].

According to the GN model, the risk of infection in a volume V, where one infective subject is present, and the initial number of viral particles is n_0_ (which can be different from zero) is given by the general formula:

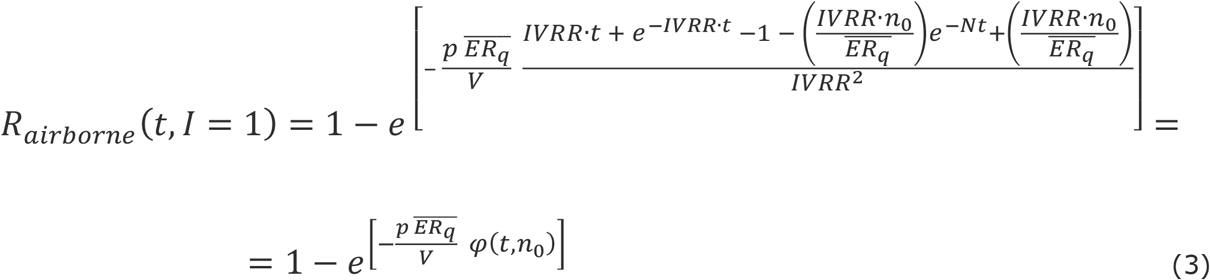

In (3) φ(t) is also a function of source and ambient parameters

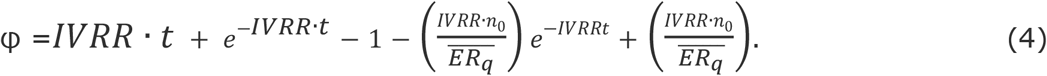

In the present model, the IVRR term in eq. (3) and (4) is dominated by natural ventilation (the AER parameter). Different factors affect natural ventilation in a classroom, such as temperature difference between the classroom and the outdoor space during windows opening, wind direction and average wind speed, as well as geometric factors such as window size and position [15]. As for thermal effects in winter conditions, a high temperature difference |*T*_*i*_ *− T*_*e*_| between indoor and outdoor temperatures is expected. In the present study we investigated a typical winter condition: indoor temperature maintained at *T*_*i*_ = +20°*C* by the school heating system and an external temperature near to 0 °C, so that |*T*_*i*_ *− T*_*e*_| = 20 °C.

This impacts the through-window ventilation flow. According to the recently revised Euronorm 16798-7:2018 [23] the single-sided airflow Q_w_ through an open windows with a given T-difference is approximately estimated by the formula (valid for moderate wind velocities): 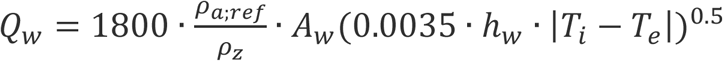 with 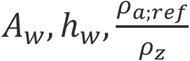 beeing the total open windows area, the windows height and the air density ratio between reference air density and density of the considered zone. This implies an AER through windows opening which becomes function of the internal-external temperature difference:

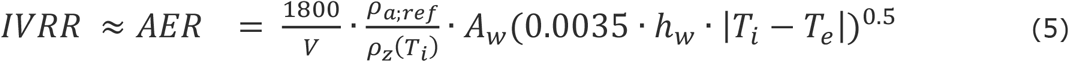

All curves were calculated for a typical classroom of volume 8 * 7 * 3 ≅ 170 m^3^, with an effective volume for aerosol diffusion of 150 m^3^. Hence, for such a standard classroom the resulting AER during windows opening results as:

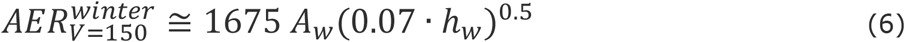

**Table 1.** Model parameters and related value ranges. In brackets the controllable parameters.

| Parameters | Description | Units | Range |
| --- | --- | --- | --- |
| $t$ | duration exposure to infection | $h$ | 0-5 |
| $t_{brk}$ | breaks duration | $min$ | [5-30] |
| $t_{lec}$ | lecture duration | $min$ | [30-100] |
| $I$ | Number of infective sources | persons | 1 |
| $S = N - I + 1$ | Number of susceptible persons | persons | 15-30 |
| $N$ | Number of students per classroom | persons | 15-30 |
| $C$ | Number of infected persons (contagions) | persons | 0-30 |
| $R=C/S$ | infection risk | - | 0-100% |
| $ER_q$ | quanta emission rate by infective source | quanta $h^{-1}$ | [5-25] |
| $p$ | average inhalation flow | $m^3h^{-1}$ | 0.6 |
| $V$ | classroom volume | $m^3$ | 170 |
| $V_{eff}$ | effective classroom volume | $m^3$ | 150 |
| $Q(t)$ | clean air inflow | $m^3h^{-1}$ | 85-1700 |
| $AER(t)_{NV,MV}$ | air exchange rate provided by natural (NV) or mechanical ventilation (MV) | $h^{-1}$ | 0-2 (NV), 0-9.5 (MV) |
| $AER_{max}$ | max air changes per hour | $h^{-1}$ | 2 (NV), 9.5 (MV) |
| $\Lambda$ | inactivation rate | $h^{-1}$ | 0.5 [28] |
| $K^{**}$ | deposition rate | $h^{-1}$ | 0-0.25 [16] |
| $IVRR(t)$ | infective virus removal rate $AER+\lambda+k$ | $h^{-1}$ | 0-2.5 (NV), 0-10 (MV) |
| $n(t)$ | viral quanta in ambient at time $t$ | quanta | 0-50 |
| $n_0$ | viral quanta at $t=0$ | quanta | $\geq 0$ |
| $f_{in}$ | mask reduction of inward viral load (susceptible person) | - | [15-30%] surgical |
| $f_{out}$ | mask reduction of outward viral load (source) | - | [45-90%] surgical |

Supplementary Fig. S1 indicates the temperature dependence of *AER*_*V*=150_ assuming two typical values of (*A*_*w*_,*h_*w*_)* in a high-school classroom where windows are partially open. It is noted that geometrical windows parameters may also change strongly in different school buildings, or even in different classrooms in the same building. Investigating specific cases, however, lies outside the scope of the present work.

To account for the effect of PPE (personal protective equipment, in this case, face masks) in reducing both the number of viral particles generated by infective subjects, and also reducing the likelihood of inhalation of viral particles by exposed subjects, we propose a modification to Eq (3) whereby the viral inhalation term *ER*_*q*_ *p/Q* is multiplied by two scaling factors:

(1-f_out_), which represents the fractional reduction of the generated viral load, and

(1-f_in_), which represents the fractional reduction of inhaled viral load, under the assumption that all subjects are wearing a mask. Eq. (3) can then be rewritten as:

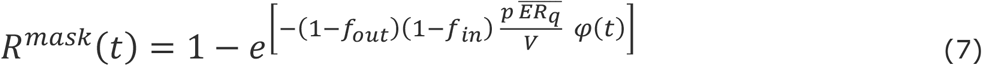

If masks are not being worn, *f*_*in*_ and *f*_*out*_ are both zero.

The extent of efficacy of face masks in reducing airborne transmission is the subject of still ongoing debate, although a general wide agreement on their importance as mitigation factor has been accepted. Some recent results [7] strongly supported the effectiveness of face masks in reducing the spread of infected aerosol droplets during exhalation, under the condition that the mask is correctly and permanently worn by both the infected and the susceptible subjects. The estimated efficacy of surgical masks in filtering the airborne viral load upon inhalation, represented by fin, varies in the available literature. Some authors (e.g. [17]) estimate the value to be close to zero, claiming that masks can only filter “large” droplets (>5µm), but more recent measurements suggest that surgical masks may actually be able to filter even “small”, i.e. sub-micrometric, droplets [24]. In the present analysis, we considered a possible range of values 0 - 0.3 for fin which is in line for surgical masks. As for the efficacy in filtering the exhaled viral load, the parameter f could have a value as high as 0.95 [7] in the case of a perfectly-adhering surgical mask worn the whole time. In a classroom environment, however, it will be difficult to ensure complete and continuous compliance over the many hours of a typical school day. For instance, a recommendation by the italian local scientific committees as of October 2020, is to wear masks for as long as possible, but to allow their occasional removal as long as social distancing is respected. Since mask filtering effectiveness varies from person to person and over the total exposure time, a rather large variation interval (from 50% to 100%) was considered for the filtering effectiveness.

The total viral load in the environment volume, in the presence of one infective subject with a rate of emission ERq > 0, is given by:

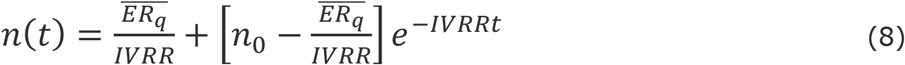

### 2.2 Cumulative risk

Applying a Wells-Riley like approach to a classroom situation where students are leaving the classroom during breaks and teacher turn classrooms, requires the underlying concept of cumulative risk. In fact, when considering the total number of infections occurred in a classroom (after a certain number of classes and breaks), the variable of practical interest is not the single risk function during a single lecture, *R*_*lec,i*_ *(t)*, (which starts from zero after each break), but the *cumulative risk R*_*c,i*_ *(t)* at the time t, which keeps into account the whole “history” of infection risk up to that point:

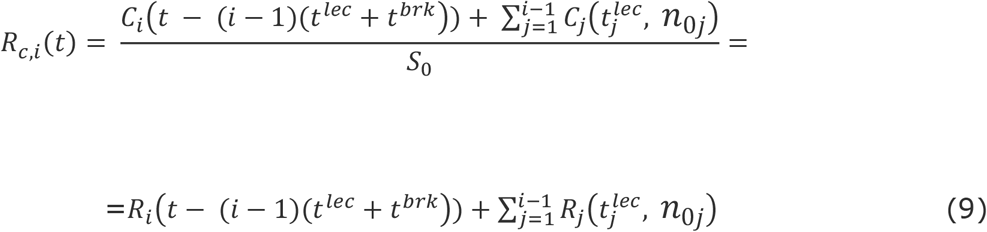

In (A1), *C*_*j*_*(t)* represents the number of infections in the previous hours and the index j spans all the “cycles” of lecture+break before the current i-lesson (j=1 to i-1, assuming cyclic lecture+breaks of duration t_lec_ + t_brk_). R_i_(t) is the infection probability during the i-cycle which is actually a dual function:

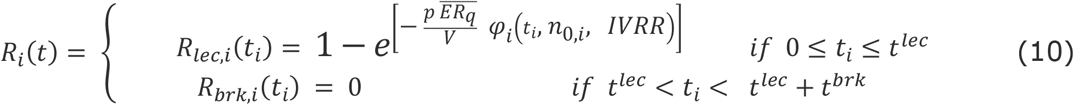

The same for the viral load in ambient:

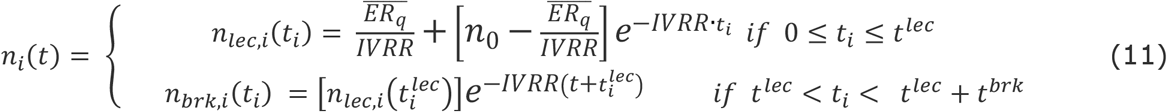

When the infective source is removed from the environment, the source parameter in equations (7) and (9) in text become zero. However, the infection risk 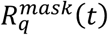 remains greater than zero, because, as a probability measure linked to the total emitted viral charge, it should keep memory of previous possible contagions. Mathematically, this is assured by the multiplicative factor φ_*i*_(*t*) in (7) which, intuitively, is non-zero because of the viral load already present in the environment after a previous lesson, even if one source left the room afterward.

Two python routines which implements equations (9-11) were developed for the infective teacher and the infective student case. For at least one infection to occur, the cumulative group risk *R*_*c*_*(t)= C(t)/S*_*0*_ must be greater than 1/ *S*_*0*_. Therefore, the condition for zero infections to occur over the total exposure time is:

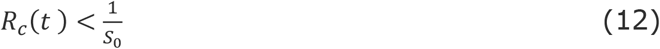

and not R_lec,i_ (t) < 1/*S*_*0*_.

It is noted that no new source should be considered within the 5h, even after a new infection occurred in that time, since any new infected person needs an incubation time of some days period before becoming infective.

### 2.3 Average emission rate for SARS-CoV2

A correct estimation for the emission rate parameter ER_q_ is the most critical assessment in the GN-Riley approach and mostly based either on a semi-empirical approach combining viral load measurements from clinical trials and fitted data from observed _out_breaks or measurements from the exhaled droplet population, which is the method followed here. Microdroplets populations in the emitted aerosol vary depending on the specific voice activity (voiced counting, whispered counting, unmodulated vocalization, breathing). The emission rate of one specific expiratory activity is described by the following analytical expression from Buonanno [18], where the index j refers to that activity (the integral formula can be well approximated with a 4 channel particle size distribution as shown by Morawksa in [25]):

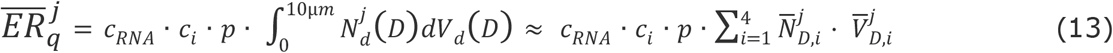

In (13) the parameter *c*_v_ is the viral load in the sputum (viral RNA copies mL^−1^) to be estimated experimentally via clinical assessments of viral loads, c_*i*_is a conversion factor having units in quanta.RNA copies^−1^ (ratio between one infective quantum and the infective dose), *p* is the pulmonary inhalation rate (m^3^ h^−1^) related to a particular k-th body-activity (resting, standing, etc). 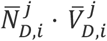 (related to the voice activity) is the product between the average droplet density and the average droplet volume (mL m^−3^) in one of the 4 channels of a typical droplet distribution expelled during voicing or breathing. For SARS-type viruses, experimental estimations of all parameters in equation (13) are reported in [18, 20, 21].

In addition, while modelling the real behaviour of a teacher or a student as a potential source one should consider a further time-average of (13). The reason is that both sources (during teaching or while attending lesson) do not behave permanently in one fixed category of vocal activity and pulmonary rate over the emission time. Therefore, the effective emission rate to be used in the model is a double averaged 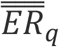 :

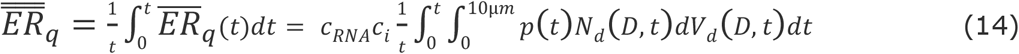

One way to estimate 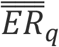 is to consider a student as a resting person and a teacher as a standing person, so that (14) can be approximated as:

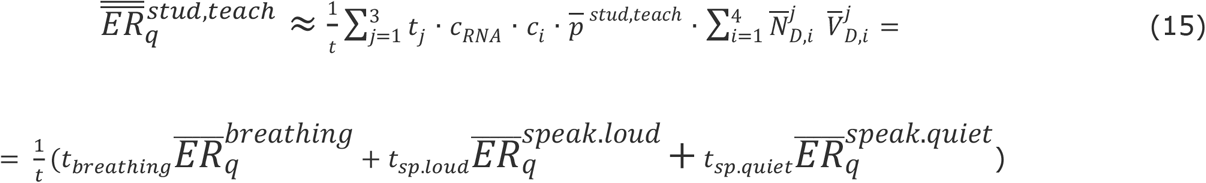

Detailed estimations of the 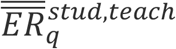used in all simulations are reported in appendix. In our model we derived values of 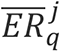 from Buonanno et al. [18] and derived 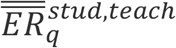 based on equation (15). Only two “voice activities” and their related effective times *t*_*j*_ were considered (breathing without speaking and with speaking). In fact, during teaching one may observe speaking periods alternated with pauses. Secondly, voice volume of a teacher may vary considerably during lesson [26]. The same for students who may put questions or comments and randomly vary their voice activity and intensity.

Even if teachers stay in a given classroom in average for two hours only, they are speaking most of this time. In primary and secondary schools, they are also speaking loudly and sometimes screaming. This makes them a much greater viral source compared to students. In the present analysis, the obtained 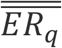 value was 32 quanta h^−1^ for an infective teacher speaking at a moderately high volume, whereas a half value of 16 quanta h^−1^ was supposed for the same subject speaking quietly through a microphone. For infective students sitting at their desk most of the time, lower values were estimated, as they were considered resting persons speaking less frequently than teachers.

### 2.3 Influence of windows opening function on the infective virus removal rate (IVRR)

Windows opening implies a periodic activation of air exchanges per hour, which are supposed to occur only during lesson breaks. Therefore, the IVRR function in (3) become a periodic rectangular wave function over the full lesson time, with peaks influenced by higher values of the air exchange rate (AER) as due to partial windows opening (2vol/h) or almost complete windows opening (up to 10 vol/h) as estimated according to EN 16798-7 [23]. In this study we explored the influence on the classroom risk function of two different cycles of the AER function. Two different ventilation cycles (long and short) were calculated for each curve (10min/50min interval/lesson and 20min/100 min interval/lesson ratio respectively). Another factor to be considered is the effective volume to be considered to dilute the aerosol viral cloud under the perfect mixing approximation. According to recent CFD simulations of aerosol cloud in classrooms [22], aerosol particles from a student source would not be diluted over the entire volume even after a transient of 300s and the viral cloud volume during the first part of the emission transient would be negligible compared to V. For these reasons an effective lower volume V_*eff*_ = 0.85 V in equation (3) and (10) was considered for the present analysis.

## 3. Results and Discussion

### 3.1 Windows opening

Airborne risk curves in a classroom of 170 m^3^ with one infective source based on the thermally extended GN-model are plotted in Figs. 3a-d in log-scale. The infective teacher case is shown in Figs. 2a and b. Figs 2c and d show the infective student case for comparison (preliminary results were presented in [13]). In both cases, however, the deviation of red curves from the reference curve (“no mitigations”) highlights the significant impact of natural ventilation alone (in the red curves, face masks were intentionally not included to isolate the net contribution of air ventilation). This reduction was about 50-60% at the lecture’s end. The additional mitigation effect of surgical masks (under the assumption they are worn by all subjects) causes a further risk mitigation of 35–45 % (depending on the effective time they are properly worn). Notably, shorter (but more frequent) breaks perform better than longer ones in all risk curves.

**Figure 1.**
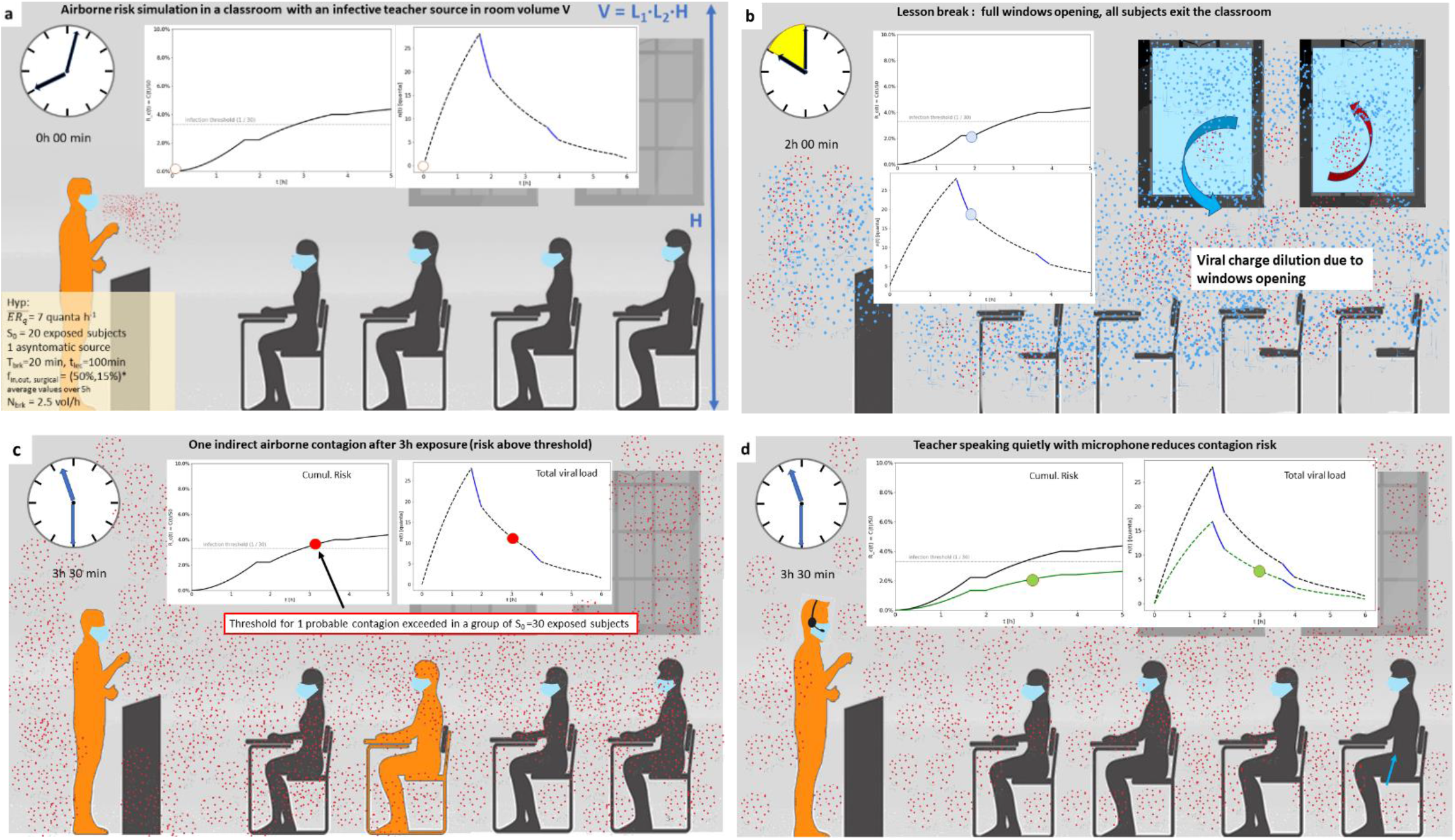
Evolution of cumulative collective risk and total viral load in a classroom of volume V with a positive teacher source (long breaks of 20 min after lectures of 100 min). All presents are supposed to wear face masks with 80% effective filter efficiency. **a)** Situation at the beginning of the lesson, white markers indicate R(0) and n(0). **b)** Air change/dilution due to windows opening after the first break. **c)** Situation after 3 hours with one probable infection. **d)** the same as in c) with less intense voicing preventing the contagion (teacher speaking through an amplified microphone).

**Figure 2.**
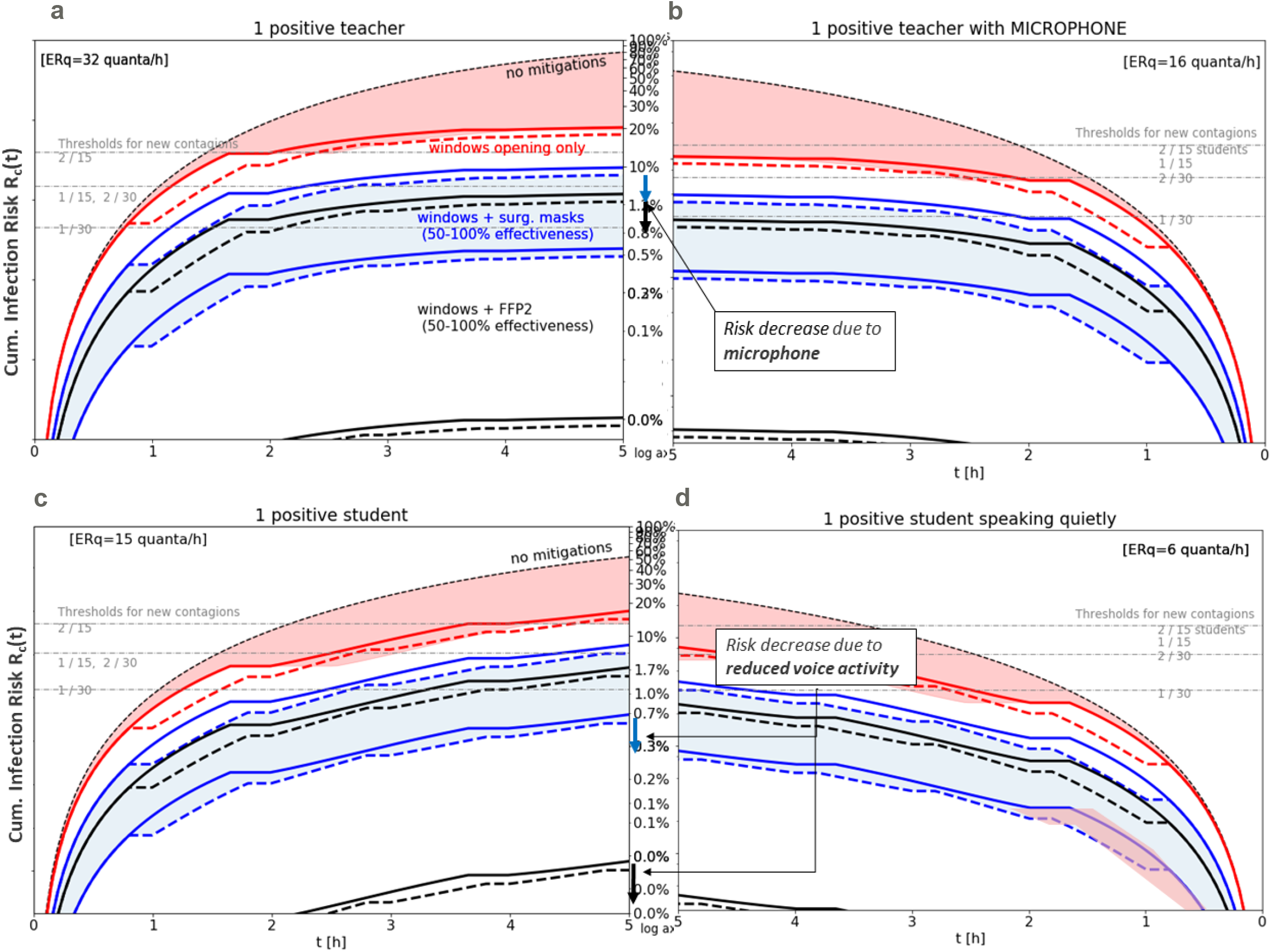
Mitigation of airborne risk in a classroom (V_eff_ = 150 m^3^) through face masks and ventilation. a) Infective teacher standing and teaching. b) Infective teacher speaking more quietly with mask + microphone + amplifier (−40% ER_q_). c) Normal infective student case. d) Infective student speaking at moderate voice volume. Partial ventilation (AER=2 vol/h) was assumed during breaks (continuous plots refer to long ventilation cycles, dashed plots to short cycles).

**Figure 3.**
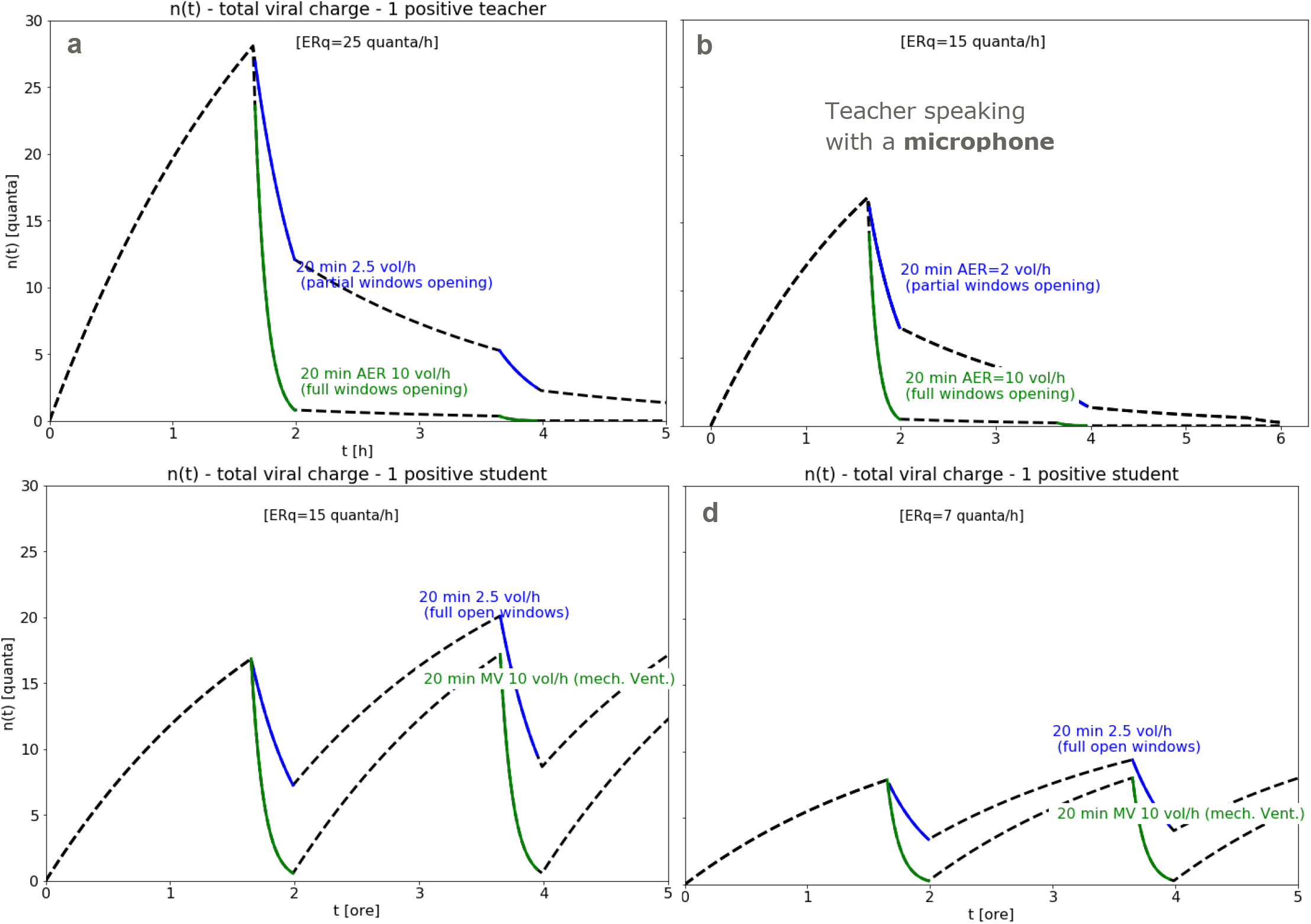
Total viral load in classroom. a) teacher speaking for 2h at normal voice volume. b) teacher speaking more quietly through a microphone + voice amplifier. Partial and full windows opening scenarios are also compared.

Teacher risk curves increase more steeply in the first 2 hours of exposure time when compared to infective student curves (Fig. 2a vs 2c). This is due to the higher average emission rate of a teaching person compared to a student sitting on a desk and speaking less frequently. In case of a microphone connected to a voice amplifier and used by a teacher, a reduced emission rate (by almost 40%) lower the risk levels for the exposed group considerably. This decrease is about −20% if considering natural ventilation as the only mitigation factor besides microphone, and up to −40% when adding the surgical masks (red and blue arrows in the middle of Fig. 2a-b). In case of a positive student source, one can still differentiate between students speaking normally and students speaking at a moderate volume (Fig. 2d). Although a microphone passed from student to student must be excluded as possible direct infection source, a more feasible scenario could be that of scholars expressly required to speak quietly. In this case, the cumulative risk levels would decrease even more: a relative delta of −50% can be observed for all curves after 5 hours exposure time. This fact can be explained with a more prolonged exposure of individuals to the infective source (5h instead of 2h) and, at the same time, a larger timespan for the mitigation effect to act (voice reduction). After the teacher has left the room, the ER_q_ in that room drops to zero, but the viral charge previously emitted by him/her will still be present for the next hours until the end of the lesson (although it will lower down after several ventilation cycles — as indicated from n(t) plots in Fig. 3). According to the GN risk model, thus, rest viral load is responsible for a further (although lower) increase of R_c_ during the next hours, even if the teacher source is no longer present.

The case of an infective student shows some important differences in the shape and final level of risk curves. Higher levels are caused by the risk still increasing after half exposure time whereas teacher curves saturate earlier to lower levels (Figs. 2c-d). This fact is eventually due to the infective student source re-entering classroom after each break and emitting until the end of the lessons. On the other hand, the one-infection thresholds are reached earlier in case of an infective teacher (blue and black curves intersecting the dotted gray lines in Fig. 2a-c and 3b-d). It is then confirmed that crowded classes with 30 students are a clearly more dangerous situation in case of an infective teacher source.

### 3.2 Reducing the voice level of infective sources

Hence, in such a case equipping the teacher with a microphone plus amplifier would be a valuable mitigation countermeasure. Moreover, due to the lowering of one-contagion-threshold, crowded classes of 30 students in limited volumes (V < 170 m^3^) should be avoided. A recommended alternative would be the splitting of the class in smaller groups alternating in face-to-face mode (Fig. 6). As a rule, keep the number of students per classroom as low as possible helps reducing the contagion risk because obviously the infection threshold lowers as N increases.

### 3.2 Classroom Volume

Airborne risk is also strongly affected by classroom volume as illustrated in Fig. 4. In classrooms of doubled volume (300 m3 instead of 150 m3) the infection probability after 5h lessons is almost the half for both cases (infective teacher and infective student). This is often the case of classrooms in historical buildings (like the Italian high school shown as an example in Fig. 5a). Here the room height may increase from standard 3m (Fig. 5b) up to 5m (Fig. 5a).

**Figure 4.**
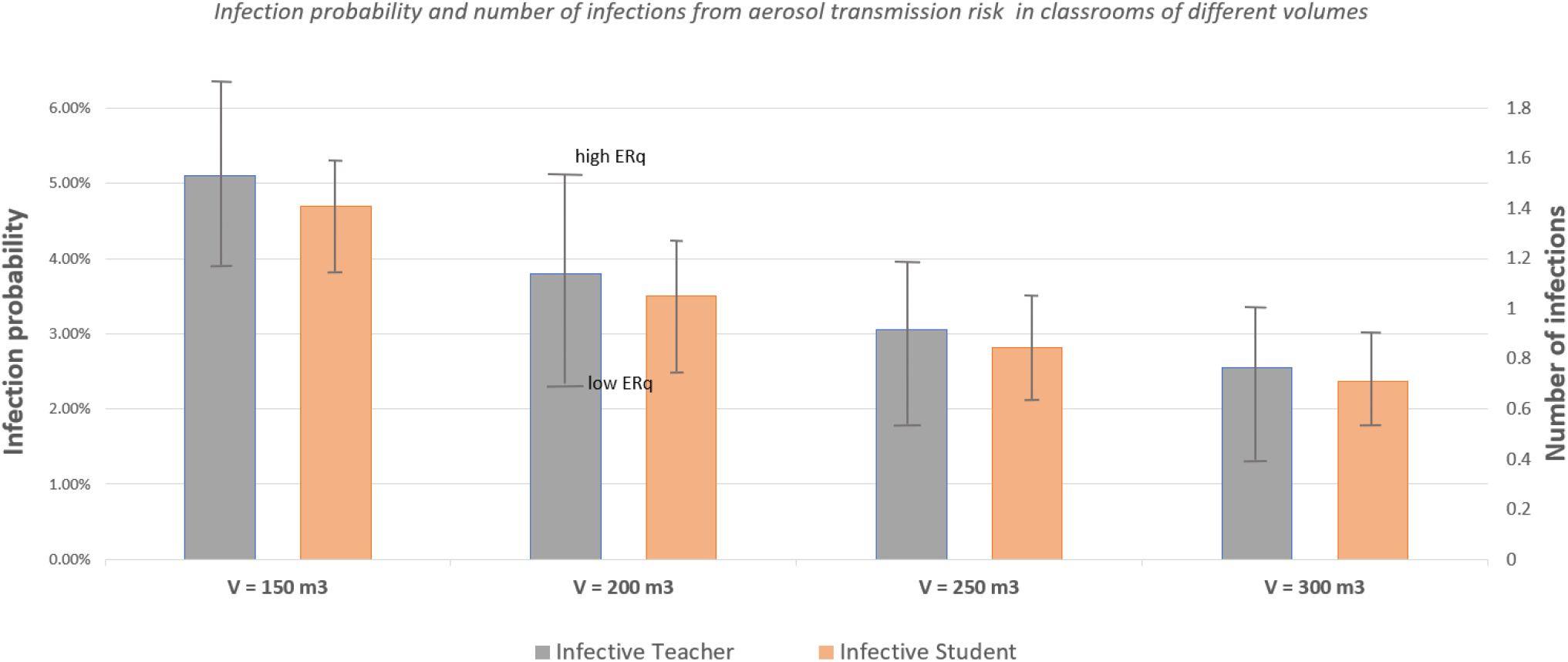
Effect of classroom volume on the infection probability calculated with the GN model (75% face mask effectiveness, surgical masks, IVRR = 5.5 vol/h at each interval). Please note the infection probability is independent of occupancy levels which only influence the number of infections C=S_0_*R_5h_.

**Figure 5.**
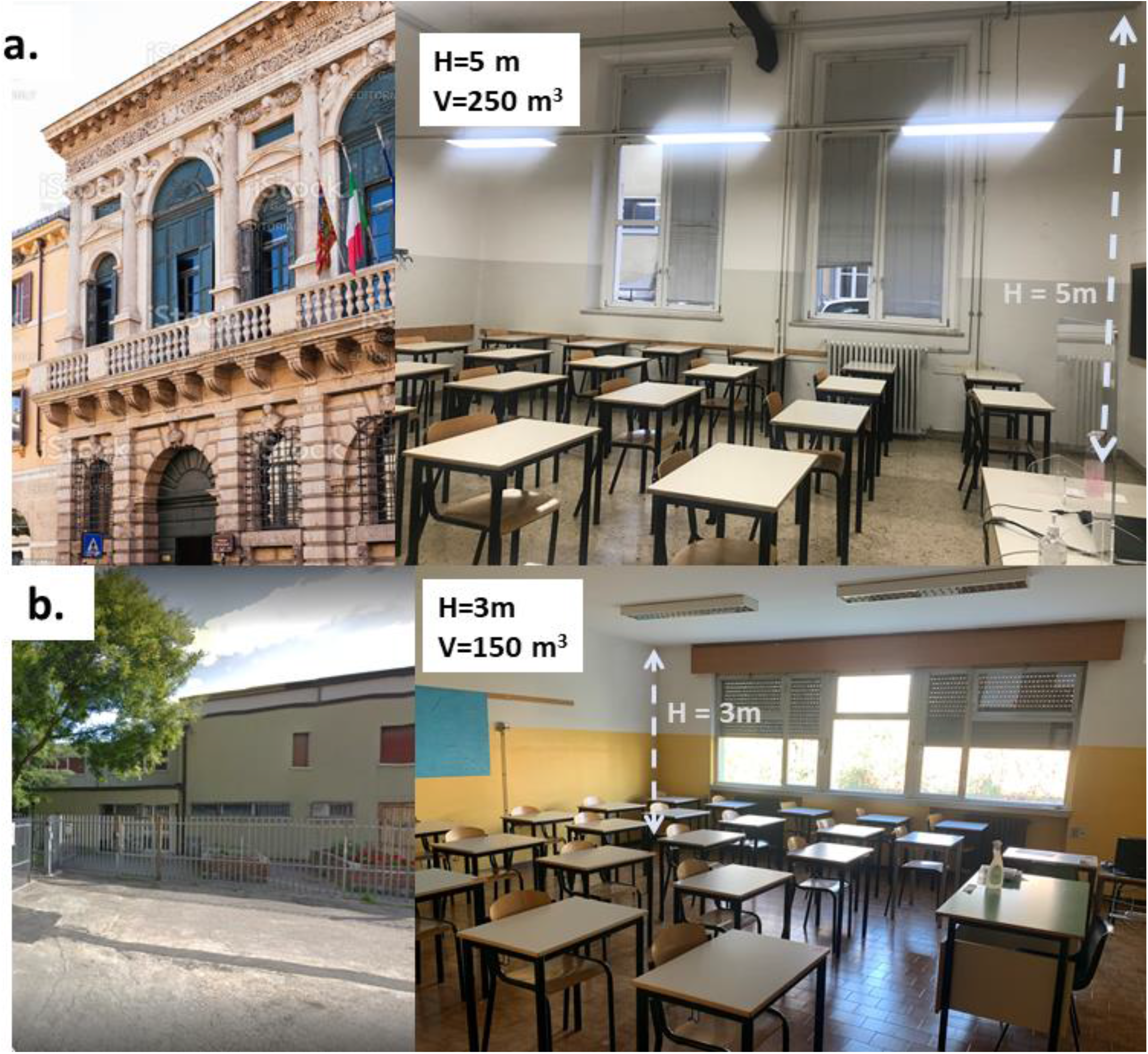
High-Schools classrooms of different heights and volumes: a. school in Italian historical building located in Verona (ITCS “Lorgna-Pindemonte”) b. standard school building also located in Verona

**Table 2.**
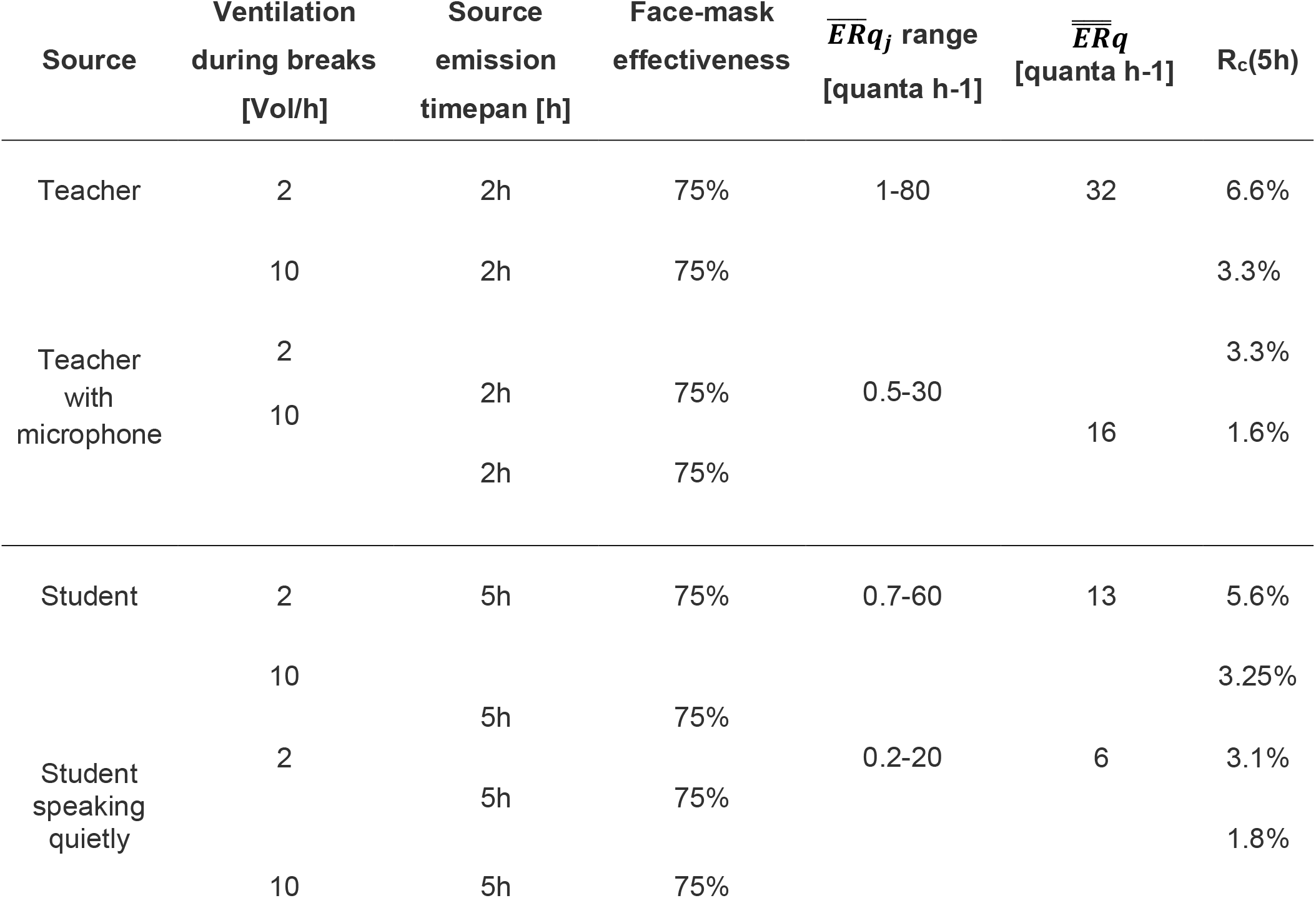
Summary of airborne infection risk values at the end of a school day in different conditions. All individuals are wearing surgical masks and classroom volume V is fixed to 150 m^3^.

## 4. Conclusions

Cumulative airborne risk is the key to understand indirect infections of SARS-CoV-2 in classrooms and possible outbreaks within a school building. The mitigation of the airborne risk in schools is linked to the main and larger goal of keeping most schools open and safe during the present pandemics pursuing a zero-infection strategy. Although the dynamic single-zone model employed here contains some approximations and also some uncertainties in the parameter estimations, the general framework is robust as it was already tested for influenza and tuberculosis and able to give clear indications for contagion risk minimization. Firstly, students and teachers are exposed in schools for relatively long time to viral aerosol and standard sanitation/ventilation cycles cannot lower the residual viral load nor the risk to zero. On the other hand, airborne risk values can be mitigated to reasonable levels by a combination of several mitigation factors. In the present situation where most schools are still not equipped with dedicated HVAC systems for controlling air ventilation and filtering at the classroom level, the regular opening of classroom windows at well-defined intervals can be an effective (although provisional) solution. Regular windows opening acts indeed as mitigation co-factor which alone almost halves the airborne risk. On the other hand, the numerical analysis confirms that only a combination of air exchange with protective masks properly worn by all exposed subjects can reduce the airborne transmission risk to acceptable safe levels.

Also splitting crowded classes of 30 students into smaller alternating groups of 15 has a dramatic beneficial effect on the collective contagion risk. This is ultimately due to the volumetric nature of the aerosol cloud (Fig. 6).

**Figure 6.**
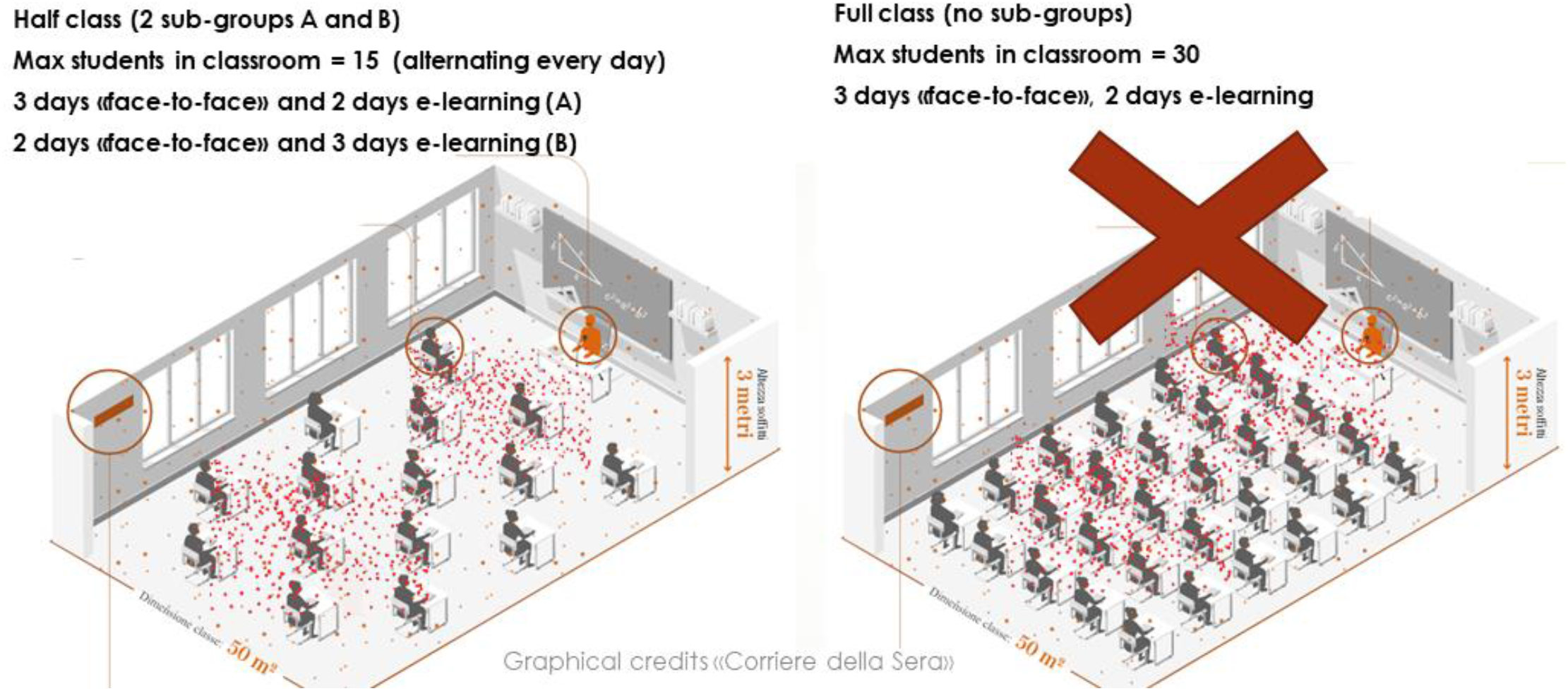
Half classes in face-to-face mode (with rotating subgroups) vs full classes on alternate days. In the latter (unrecommended) case one may note the increased number of students in contact with the viral cloud emitted by the infective source

Concerning more specific countermeasures, it has been shown that equipping teachers with microphone and voice amplifier as well as requesting students to speak quietly during lessons could be effective and feasible mitigation factors which contribute to keep the airborne risk levels below safety thresholds.

Combining all these factors, one could achieve the desired goal of collective risk below the one infection threshold in a classroom during the full exposure time (5h). However, it is remarked that a natural ventilation strategy was suggested as a compromise emergency solution for the vast majority of schools still not equipped with HVAC systems. For the middle and long-term future, equipping schools with dedicated HVAC systems remain a preferrable option for what concerns air quality, energy efficiency, thermal comfort and ventilation control. The 3D nature of aerosol diffusion also suggests a revision of safety regulation and social distancing in schools which should consider a new volumetric approach. For instance, while linear social distancing (varying from 1m to 2m in different EU countries) could be maintained, a higher minimum value for the classroom height (and therefore a larger minimum classroom volume) should be considered in regulations. Many historical buildings, for instance, already have much higher classroom volumes due to higher internal walls (up to 50-80% higher than in standard school buildings). A revision of social distancing norms in schools (currently based on linear distancing) is also urgently required. Classrooms compliant for social distancing but small in height, should be furtherly checked to guarantee a minimum height (and volume). If a relatively small volume would cause the risk function to increase above the contagion thresholds, one of the illustrated mitigations should be adopted. To this regard, while schools in historical buildings lack the possibility to install centralized HVAC systems, they could more easily comply the volumetric requirement in individual classrooms.

We conclude with a statistical remark. As previously noted, in regions where the likelihood of one or more asymptomatic source seems particularly high or to increase steeply, a class splitting strategy would be highly recommended, particularly for large groups (≥ 25 students). However, to be more precise we should base this decision on the probability to have one or more asymptomatic sources in a classroom (the present study already assumes one asymptomatic source). Therefore there is a need to solve an open statistical problem: the estimation of the local probability function p_local_(I ≥1 | N_age_) of having at least one asymptomatic source (but they could be even more than one) in a random ensemble of N students of given age located in a certain region, given the local epidemic situation in that particular area, city or district.

## Data Availability

theoretical/computational work based on running python scripts. Python scripts are available upon request

## APPENDIX

**Table A1.** Estimations of 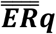 values adopted in simulations

| | | $ER_{q,min}$ | $ER_{q,max}$ | $\overline{ERq}_j$ | $t_{br}$ | $t_{sp,low}$ | $t_{sp\_normal}$ | $\overline{ERq}$ | $\pm\sigma$ |
| --- | --- | --- | --- | --- | --- | --- | --- | --- | --- |
| Voice |  |  |  |  |  |  |  |  |  |
| Activity |  | (quanta/h) |  |  | (% time) |  |  | (quanta/h) |  |
| Teacher | normal | 1 | 80 | 40,5 | 15 | 10 | 75 | 32,1 | $\pm 6,2$ |
|  | low | 0,5 | 30 | 15,25 |  |  |  |  |  |
|  | breathing | 0,1 | 2 | 1,05 |  |  |  |  |  |
| Teacher +<br>microphone | normal | 1 | 80 | 40,5 | 15 | 75 | 10 | 15,4 | $\pm 3,0$ |
|  | low | 0,5 | 30 | 15,25 |  |  |  |  |  |
|  | breathing | 0,1 | 2 | 1,05 |  |  |  |  |  |
| Student | normal | 0,7 | 70 | 35,35 | 65 | 0 | 35 | 13,1 | $\pm 2,5$ |
|  | low | 0,2 | 20 | 10,1 |  |  |  |  |  |
|  | breathing | 0,1 | 2 | 1,05 |  |  |  |  |  |
| Student<br>speaking<br>quietly | normal | 0,7 | 70 | 35,35 | 65 | 35 | 0 | 6,0 | $\pm 1,2$ |
|  | low | 0,2 | 30 | 15,1 |  |  |  |  |  |
|  | breathing | 0,1 | 2 | 1,05 |  |  |  |  |  |

## Supplementary Figures

**Figure S1.**
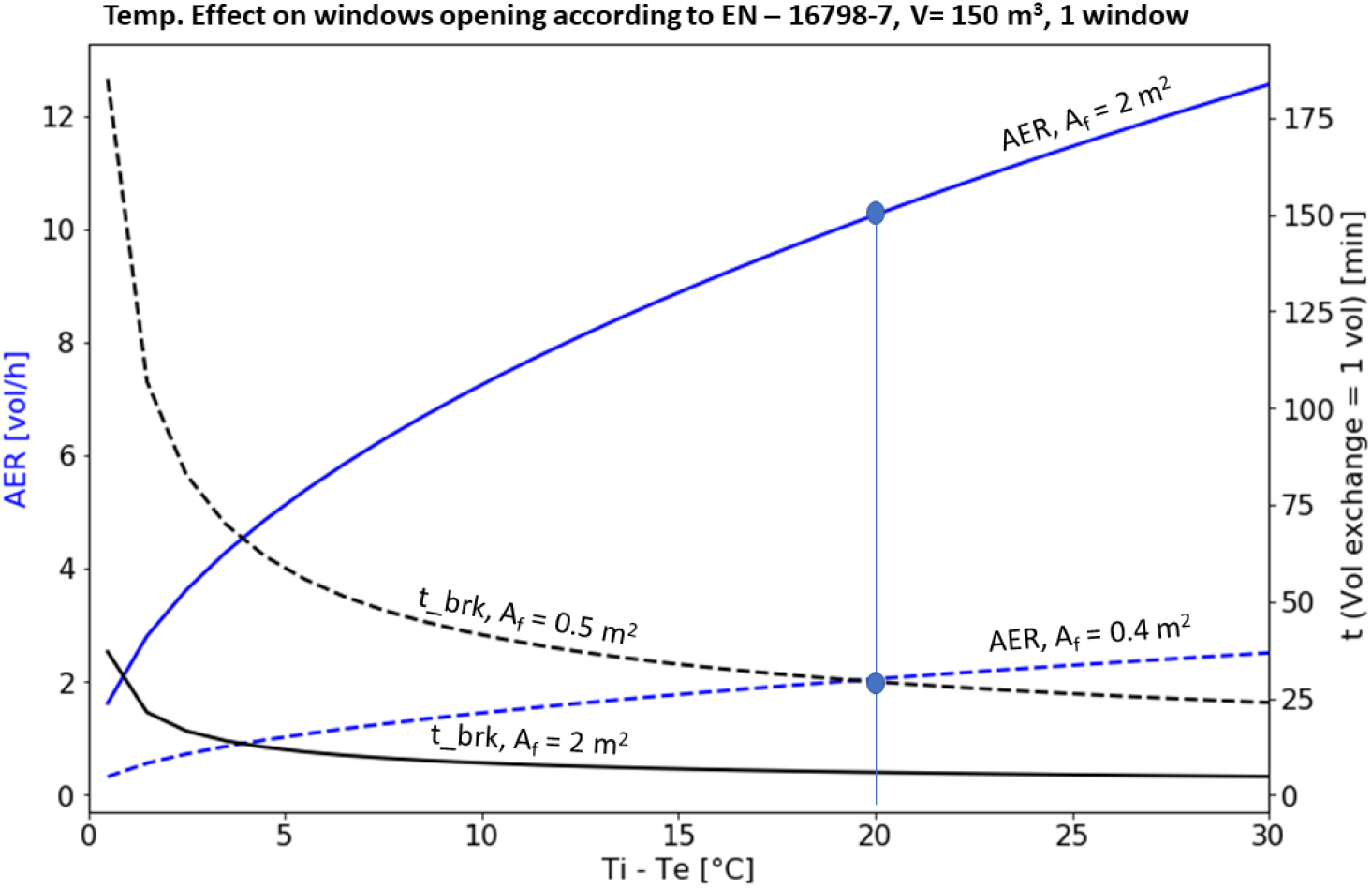
Effect of indoor/outdoor temperature difference on the air-exchange-rate by windows opening (blue curves) in a typical classroom according to the EN 16798-7 (single-sided ventilation) [23]. Marked blue dots indicate AER values used in simulations at Ti – Te = 20°C. Times for one exchanged volume are also shown.

## Notes

### Competing Interest Statement

The authors have declared no competing interest.

### Funding Statement

no external funding was received

### Summary of Updates

improved text and added 2 figures

